# Viral Kinetics of Severe Acute Respiratory Syndrome Coronavirus 2 (SARS-CoV-2) Omicron Infection in mRNA-Vaccinated Individuals Treated and Not Treated with Nirmatrelvir-Ritonavir

**DOI:** 10.1101/2022.08.04.22278378

**Authors:** Eric Y. Dai, Kannon A. Lee, Audrey B. Nathanson, Ariana T. Leonelli, Brittany A. Petros, Taylor Brock-Fisher, Sabrina T. Dobbins, Bronwyn L. MacInnis, Amelia Capone, Nancy Littlehale, Julie Boucau, Caitlin Marino, Amy K. Barczak, Pardis C. Sabeti, Michael Springer, Kathryn E. Stephenson

## Abstract

We measured viral kinetics of SARS-CoV-2 Omicron infection in 36 mRNA-vaccinated individuals, 11 of whom were treated with nirmatrelvir-ritonavir (NMV-r). We found that NMV-r was associated with greater incidence of viral rebound compared to no treatment. For those that did not rebound, NMV-r significantly reduced time to PCR conversion.

## BACKGROUND

The protease inhibitor nirmatrelvir-ritonavir (NMV-r) decreases the risk of severe COVID-19 (coronavirus disease 2019) in unvaccinated high-risk outpatients with SARS-CoV-2 (severe acute respiratory syndrome coronavirus 2) infection^1,2^, but has not shown clinical benefit in low-risk and/or vaccinated individuals^3^. NMV-r accelerates time to viral suppression in a range of populations^1,3,4^, suggesting that there may be a public health benefit of NMV-r treatment if reduced viral shedding translated to decreased transmission. However, there are concerns that cases of viral rebound following NMV-r cessation with prolonged viral shedding would negate any such public health benefit. Such cases have been described in mRNA-vaccinated individuals infected with the Omicron variant, where viral rebound was characterized by recurrence of symptoms, high viral load, a long duration to re-suppression of virus, and the presence of culturable virus, though not severe disease or impaired SARS-CoV-2 specific immune responses^5-8^.

The data on the incidence of viral rebound in treated and untreated populations is mixed, primarily due to varying definitions of rebound, methods of ascertaining events, and patient populations^1,4,9-11^. In the largest dataset of viral kinetics in untreated mRNA-vaccinated individuals infected with the Omicron variant, viral rebound (defined with PCR monitoring) occurred in 6% of 494 infections^10^. To date, however, there is no direct comparison of the incidence of viral rebound in NMV-r-treated and untreated individuals in a similar mRNA-vaccinated population.

To address this knowledge gap, we conducted a prospective observational study of 36 individuals newly diagnosed with SARS-CoV-2 infection in Massachusetts, United States between March and May 2022, 11 of whom had initiated therapy with NMV-r at the time of enrollment. We collected anterior nares samples daily for at least 14 days for viral load quantification and whole genome viral sequencing; 1 additional swab was collected at enrollment for viral culture. All participants completed questionnaires reporting demographics, past medical history, COVID-19 vaccination status, and symptoms.

## METHODS

### Study Participants

Individuals were eligible for participation if greater than 2 years old and diagnosed with SARS-CoV-2 infection by rapid antigen test or polymerase chain reaction (PCR) within 7 days of enrollment. The protocol was approved by the Beth Israel Deaconess Medical Center institutional review board. Study staff reviewed study requirements verbally with eligible individuals and obtained and documented verbal consent, and verbal assent for children as applicable.

### Endpoint Assessments

Detailed methods with citations are described in the supplement. Participants self-collected (unsupervised) anterior nasal swab specimens at home for a minimum of 14 days. Virus was quantified with the Quaeris SARS-CoV-2 Assay, a real-time reverse transcription polymerase chain reaction (rRT -PCR) test. Cycle threshold (Ct) values <35 were considered positive. Viral rebound was defined as at least 2 negative (Ct≥35) PCR results followed by at least 2 positive (Ct<35) results. SARS-CoV-2 genomes were sequenced and aligned to the Wuhan-Hu-1 reference genome. Single-nucleotide variations (SNV) were called relative to Wuhan-Hu-1 and visualized relative to the BA.2 reference, as all 8 sequenced samples were of the BA.2 lineage and its descendants. Viral culture was performed from an enrollment swab and assessed semi-quantitatively by median tissue culture infectious dose assay (TCID50).

## RESULTS

36 participants were enrolled between March 4, 2022 and May 25, 2022. The median age was 44 years (range 25-48) in the NMV-r group, and 16 years (range 5-66) in the untreated group (Supplemental Table 1). Participants in the NMV-r group were more likely than in the untreated group to have comorbidities, to have ever smoked/vaped, and to be overweight/obese. There was a median of 3 prior COVID-19 vaccine doses in both groups; all participants had received at least one mRNA vaccine. All sequenced viruses were BA.2 or a sub-lineage.

One (1) out of 25 individuals in the untreated group (4%) had virologic rebound compared to 3 out of 11 (27%) in the NMV-r group (p=.04, Chi-square test, Figure 1A-B). Among individuals who did not have viral rebound, the duration of PCR positivity from initial positive test (diagnosis) to last positive PCR was significantly shorter in the NMV-r group vs. the untreated group (median 3.5 vs. 7 days, p=.0006 by Mann-Whitney test, Figure 1C). This difference in time from initial diagnosis to last positive PCR was no longer significant when including rebound cases where last positive test was counted at end of rebound (Supplemental Table 4). Among those with viral rebound following NMV-r, the median time to rebound from end of initial infection (first negative test) was 5 days (range 2-7) and the median peak virus level during rebound was Ct=19 (range 24.8 to 17.2). In the single case of rebound without treatment, time to rebound was 5 days and peak virus level was Ct=30.3. There was no difference in peak detected virus level during the initial infection period between those treated with NMV-r who had virologic rebound, and those that did not (median Ct=29 vs. Ct=27.5, Figure 1D), though there was a suggestion of a longer time to undetectable viral load (Ct≥40) (median 5 vs. 3 days, Figure 1E). Within the NMV-r group, live virus was cultured from 1/3 samples (collected at enrollment) among rebounders compared to 0/4 samples from non-rebounders (Figure 1F). Baseline symptoms and other characteristics were similar between rebounders and non-rebounders in the NMV-r group (except that the only immunocompromised individual in cohort was in the rebound group, Supplemental Table 2). All NMV-r treated participants had received at least 3 mRNA vaccinations. One case of NMV-associated rebound was asymptomatic.

**Figure 1.**
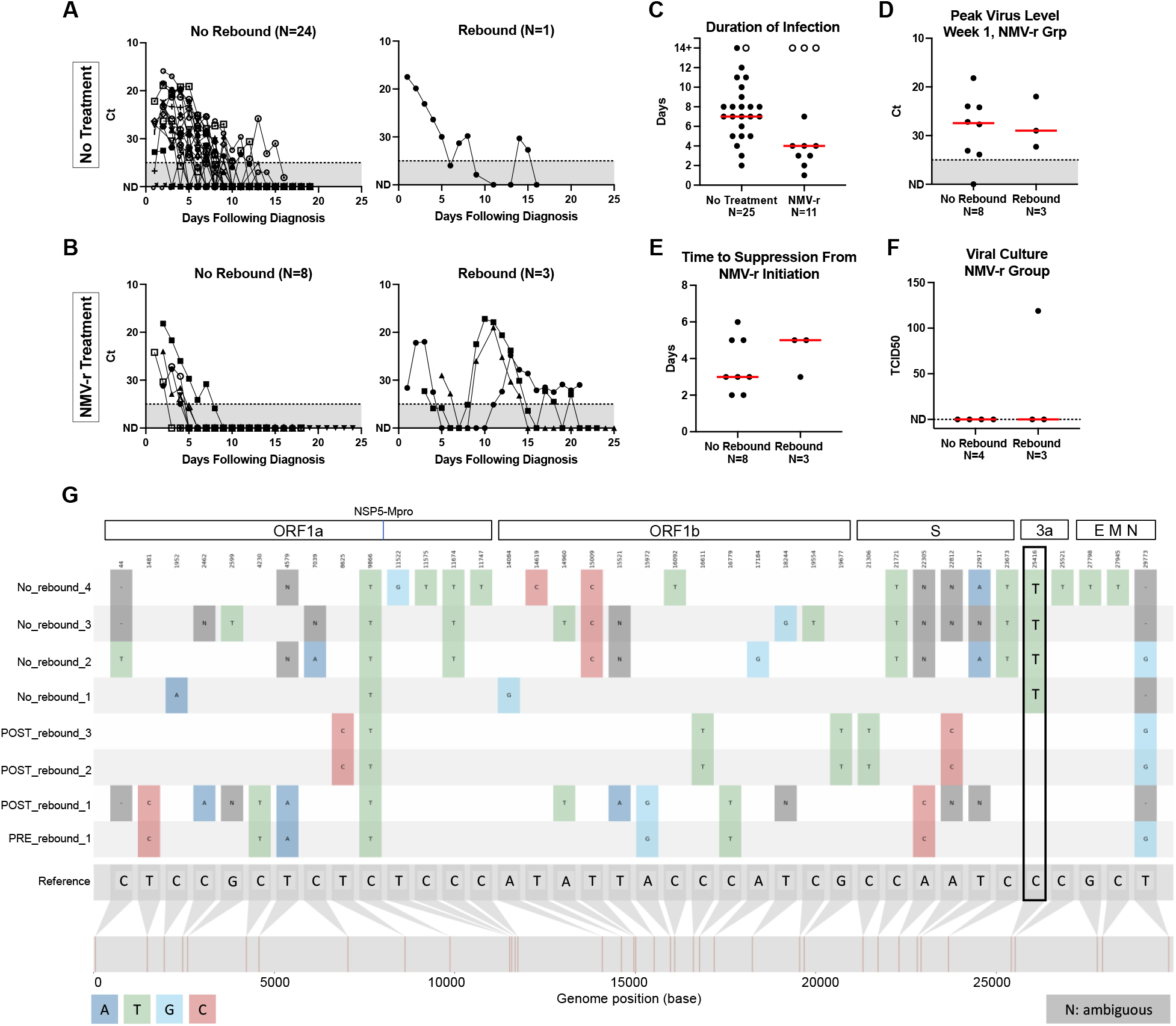
Viral kinetics of SARS-CoV-2 infection in participants treated and not treated with NMV-r. (A-B) The magnitude of virus is plotted on the *y* axis as Cycle threshold (Ct) value; numbers are plotted inversely to show that a lower Ct value signifies a higher amount of virus. Days following diagnosis (first positive test) are plotted on the *x* axis. Grey shading indicates the Ct threshold (35) for negative/positive determination. Left panels depict non-rebounders. Right panels depict rebounders. Participants who did not receive treatment (N=25) are plotted in (A). Participants who received NMV-r treatment are plotted in (B). Dotted line indicates Ct=35, the threshold for positivity. ND=not detected, plotted as Ct=40. (C) Duration of viral shedding (time to negative PCR up to at least 14 days) for no treatment and NMV-r treatment groups. Open circles indicate rebounders. (D) Peak virus level (lowest Ct) detected during initial infection period in NMV-r treatment group plotted by rebound status. (E) Time to suppression (days to negative PCR, Ct<40) following initiation of NMV-r plotted by rebound status. (F) SARS-CoV-2 live virus culture titer (TCID50) of nasal samples from day of enrollment in NMV-r treatment group plotted by rebound status. ND=not detected. Solid red bars in (C-F) indicate median values. (G) Plot of SARS-CoV-2 whole genome alignment, relative to the BA.2 reference sequence, for viruses in NMV-r treatment group plotted by rebound status (“no rebound”, “pre-rebound”, and “post-rebound”). Single nucleotide variations from reference sequence are noted in color. Participant numbering for sequence analysis (e.g. “Rebound Participant #1” etc.) is arbitrary for publication purposes and does not match study identification numbers or the lettering system used in Supplemental Table 2.

Viral isolates from initial and rebound infections were sequenced from 4 non-rebounders and 3 rebounders (both pre- and post-rebound) in the NMV-r group. One single nucleotide variation (C25416T) was detected in viruses from initial infection that was unique to non-rebounders compared to rebounders, who all possessed the ancestral allele both before and following NMV-r treatment (Figure 1G; pre-rebound samples with poorer sequence quality included in Supplemental Figure 1), Of note, 2 rebounders were from the same household and thus epidemiologically and genomically linked. This nucleotide variation (C25416T) was present on the accessory gene ORF3a. No mutations associated with NMV-r resistance were identified in initial or rebound viruses.

## DISCUSSION

In this small prospective observational study of mRNA-vaccinated individuals, we found that viral rebound was observed significantly more often following NMV-r treatment compared to no treatment. In the 3 cases of rebound following NMV-r, viral kinetics during the period of rebound were similar to that observed in acute infection, with an abrupt rise to high peak levels (median Ct=19) and 6-12 days to viral clearance. In the rebound case following no treatment, viral kinetics were more akin to a so-called “blip”, e.g., 2 days of positive values with peak Ct=30.3.

The baseline incidence of rebound in the untreated group was 4% when rebound was defined as at least 2 negative (Ct≥35) PCR results followed by at least 2 positive (Ct<35) results. Estimates of the incidence of rebound vary widely across the literature because of differences in methodology^4,9-11^. Our result is consistent with the 6% rate reported in a prospective observational cohort of mRNA-vaccinated individuals infected with Omicron, where viral rebound was defined using similar PCR criteria. Among the 11 individuals who were treated with NMV-r in our study, we observed viral rebound in 3 cases (27%). Given the small number of events, a precise estimate of the true incidence of NMV-r viral rebound in mRNA-vaccinated individuals cannot be made from this data.

For those that did not have viral rebound, NMV-r treatment significantly reduced the duration of infection (median 7 vs. 3.5 days). However, when including rebounders in the analysis, the difference in time from diagnosis to last negative PCR (including rebound period) was no longer significant. Further studies with larger sample sizes and/or in dense transmission units (e.g., households) are required to determine if treating low-risk/vaccinated populations with NMV-r can prevent secondary SARS-CoV-2 transmission.

It is unclear why rebound occurs in some individuals following NMV-r treatment and not in others. Our data suggests that there may be differences in the early dynamics of infection that are unmasked by treatment. In comparison to non-rebounders, infection in rebounders took longer to suppress to undetectable (Ct≥40) with NMV-r (median 5 vs. 3 days) and viruses from initial infection were more likely to grow in live culture (1/3 vs. 0/4 cultures). Further, we found that sequenced viruses in the rebound group lacked a single nucleotide variation in Orf3a, an accessory gene that is not associated with NMV-r resistance but when deleted, significantly attenuates replicative capacity in animal models^12^. However, given that viruses from 2 of the NMV-r rebounders are epidemiologically linked, the lack of this mutation may not be meaningful.

NMV-r remains a critical tool for treating outpatient SARS-CoV-2 infection in patients who are at high risk for progressing to severe disease, e.g., individuals with comorbidities who are unvaccinated or may have diminished vaccine-induced immunity. Optimal deployment of NMV-r as an antiviral agent will require defining the effect in other populations, identifying the host and viral factors that put treated individuals at risk for rebound, and determining the consequences of rebound on forward transmission.

## Data Availability

All data produced in the present study are available upon reasonable request to the authors.

## ACKNOWLEDGEMENTS

We thank the participants and staff at the Center for Virology and Vaccine Research Clinical Trials Unit, the Harvard Catalyst Clinical Research Center, and the Beth Israel Deaconess Primary Care—Chelsea Clinic.

## SUPPLEMENTAL INFORMATION

### Supplemental Methods

#### Viral Load Quantification

Viral load quantification was measured with the Quaeris SARS-CoV-2 Assay, a real-time reverse transcription polymerase chain reaction (rRT -PCR) test using the Luna Probe One-Step RT-qPCR Kit (No ROX) [NEB E3007]^1^. The SARS-CoV-2 primer and probe set is designed to detect RNA from the SARS-CoV-2 N1 and RdRP genes and the human RNase P gene in nasal specimens from suspected patients. The Quaeris assay employs a RNaseP internal control to determine that the self-collection resulted in a sample of appropriate quality and quantity of RNA. When received by the laboratory, samples are first rehydrated with 300 µl phosphate buffered saline (PBS), then inactivated at 65ºC and subsequently used directly as input for the Quaeris assay. There is no extraction step. rRT-PCR is performed on an Applied Biosystem Quantstudio 7 instrument (software version 1.7). Liquid handling is automated using either the Tecan Fluent 1080, or the Hamilton Star, or the Multidrop combi dispenser. Cycle threshold is reported for N1 gene.

#### SARS-CoV-2 Whole Genome Sequencing

Sequencing of viral isolates from initial and rebound infections was attempted for 8 unique participants in the NMV-r group (5 non-rebounders, and 3 rebounders, pre- and post-rebound). Viral RNA was extracted from anterior nares swabs using the KingFisher Flex System with the MagMAX MirVana Total RNA Isolation Kit. Illumina sequencing libraries were constructed using the ARTIC v4.1 multiplexed primer set as previously described^2^ and sequenced on an Illumina NextSeq instrument. The viral-ngs pipeline (https://github.com/broadinstitute/viral-pipelines) was used to demultiplex reads, remove adaptor and contaminant sequences, deplete human reads, and assemble genomes to the reference sequence NC_045512.2. Complete genomes (> 24000 unambiguous bp in assembled genome length) were assigned Pango lineages using Nextclade^3^ (https://clades.nextstrain.org). Visualization was prepared using snipit (https://github.com/aineniamh/snipit) to label single nucleotide variants relative to the BA.2 reference sequence (GISAID ID: EPI_ISL_8128463). Notably, the pre-rebound samples for Rebound Participants #2 and #3 were of poorer quality (3053 and 20261 unambiguous bases, respectively), but were included to demonstrate the presence of the ancestral allele before, and thus not in response to selective pressures from, drug treatment in all cases where the base was unambiguously resolved. Participant numbering for sequence analysis (e.g. “Rebound Participant #1” etc.) is arbitrary for publication purposes and does not match study identification numbers or the lettering system used in Supplemental Table 2.

#### SARS-CoV-2 Culture

We performed viral culture in the BSL3 laboratory of the Ragon Institute of Massachusetts General Hospital (MGH), Massachusetts Institute of Technology (MIT), and Harvard. Viral culture was assessed semi-quantitatively by median tissue culture infectious dose assay (TCID50) as previously reported^4^. In brief, viral swabs for culture were placed in viral transport media for storage and transport. Viral transport media was filtered through a 0.65µm filter; then used to inoculate Vero-E6 cells in serial dilutions in a 96-well format. Wells were observed with a light microscope on day 7 post-infection, and wells demonstrating CPE were scored as positive.

#### Statistical Analysis

Viral rebound was defined as at least 2 negative (Ct≥35) PCR results followed by at least 2 positive (Ct<35) results. These criteria were chosen to enhance comparability with data in Li et al.^5^ and Hay et al^6^. The incidence of viral rebound was compared between treated and untreated participants using Chi-square test. Duration of infection was calculated as days from initial positive test (diagnosis) to last positive PCR (Ct<35). Since follow up was variable beyond 14 days, positive values after 14 days was calculated as 14 days. Median duration of infection was compared between treated and untreated participants using Mann-Whitney test, both with and without rebound cases.

## Supplemental Tables

**Supplemental Table 1.**
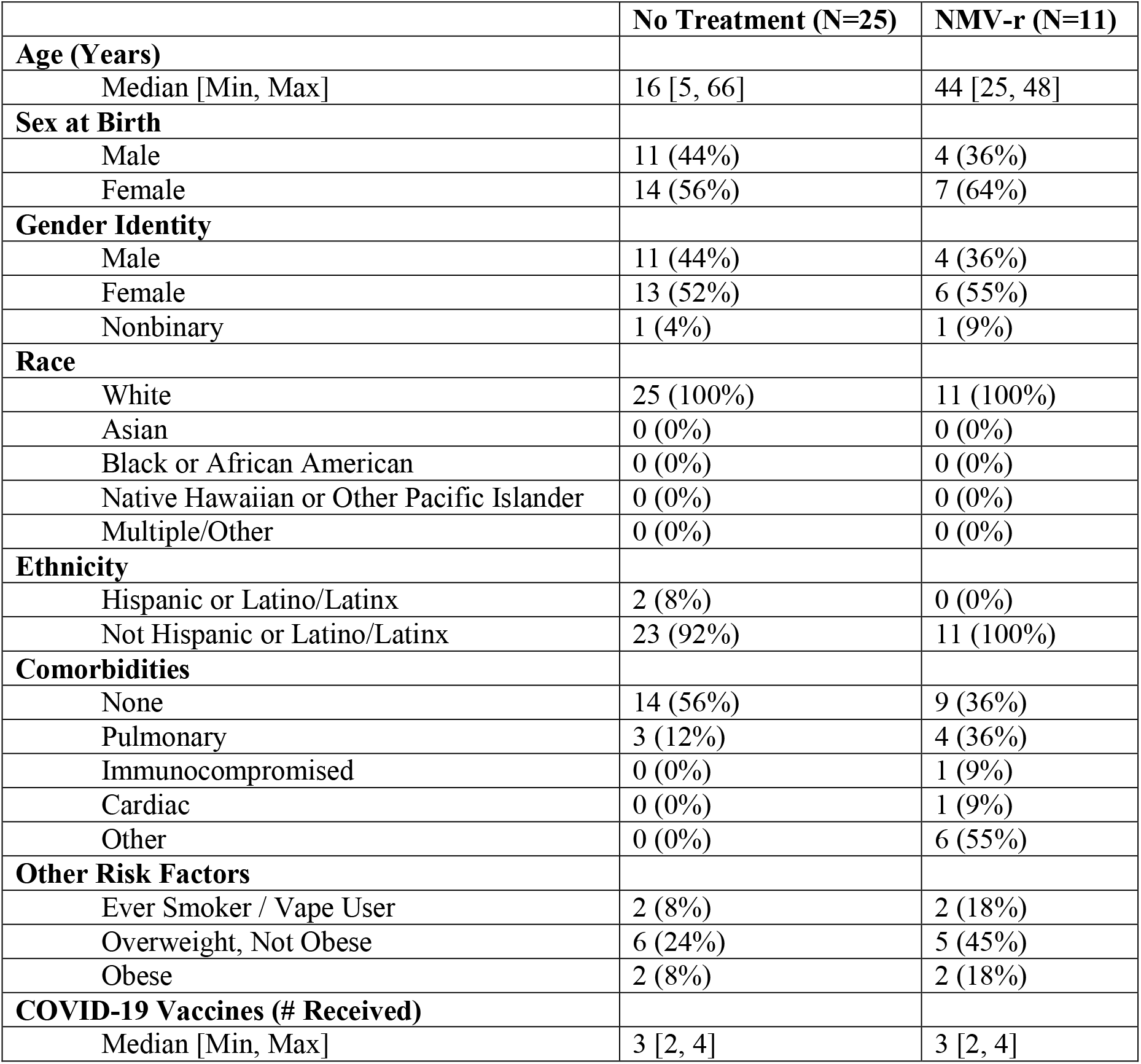
Baseline Characteristics

|  | No Treatment (N=25) | NMV-r (N=11) |
| --- | --- | --- |
| **Age (Years)** |  |  |
| Median [Min, Max] | 16 [5, 66] | 44 [25, 48] |
| **Sex at Birth** |  |  |
| Male | 11 (44%) | 4 (36%) |
| Female | 14 (56%) | 7 (64%) |
| **Gender Identity** |  |  |
| Male | 11 (44%) | 4 (36%) |
| Female | 13 (52%) | 6 (55%) |
| Nonbinary | 1 (4%) | 1 (9%) |
| **Race** |  |  |
| White | 25 (100%) | 11 (100%) |
| Asian | 0 (0%) | 0 (0%) |
| Black or African American | 0 (0%) | 0 (0%) |
| Native Hawaiian or Other Pacific Islander | 0 (0%) | 0 (0%) |
| Multiple/Other | 0 (0%) | 0 (0%) |
| **Ethnicity** |  |  |
| Hispanic or Latino/Latinx | 2 (8%) | 0 (0%) |
| Not Hispanic or Latino/Latinx | 23 (92%) | 11 (100%) |
| **Comorbidities** |  |  |
| None | 14 (56%) | 9 (36%) |
| Pulmonary | 3 (12%) | 4 (36%) |
| Immunocompromised | 0 (0%) | 1 (9%) |
| Cardiac | 0 (0%) | 1 (9%) |
| Other | 0 (0%) | 6 (55%) |
| **Other Risk Factors** |  |  |
| Ever Smoker / Vape User | 2 (8%) | 2 (18%) |
| Overweight, Not Obese | 6 (24%) | 5 (45%) |
| Obese | 2 (8%) | 2 (18%) |
| **COVID-19 Vaccines (# Received)** |  |  |
| Median [Min, Max] | 3 [2, 4] | 3 [2, 4] |

**Supplemental Table 2.** Detailed description of participants in NMV-r treatment group by rebound status

| No Rebound (N=8) |  |  |  |  |  |  |  |  |
| --- | --- | --- | --- | --- | --- | --- | --- | --- |
|  | Age Range | Sex at Birth | Risk Factors | Prior Infection | COVID-19 Vaccines | Days from Vaccination | Acute Symptoms [Max Duration] | Rebound Symptoms |
| A* | 21-25 | Female | Diabetes | No | Pfizer x 3 | 164 | Cough, sore throat, fatigue [3 days] | NA |
| B | 46-50 | Male | Former/Current Smoker<br>BMI>30 | No | Pfizer x 3 | 151 | Cough, sore throat, fever (99) [6 days] | NA |
| C | 31-35 | Female | Asthma<br>BMI>25 | No | Pfizer x 3 | 177 | Runny nose, decreased taste, sneezing, abdominal pain, joint pain, fatigue, cough, sore throat, chills, fever (101.5), headache, dizziness, muscle aches, diarrhea [8 days] | NA |
| D | 46-50 | Male | Asthma<br>BMI>25 | No | Moderna x 2<br>Pfizer x 1 | 162 | Runny nose, sneezing, fatigue, cough, shortness of breath, chills, fever (not measured), muscle aches [3 days] | NA |
| E | 26-30 | Male | None | No | Pfizer x 3 | 161 | Fatigue, cough, sore throat, chills, fever (99), chest pain, headache [4 days] |  |
| F | 46-50 | Female | Former/Current Smoker | No | Pfizer x 2<br>Moderna x 1 | 182 | Ear pain, stuffy nose, sneezing, no appetite, abdominal pain, joint pain, fatigue, cough, shortness of breath, sore throat, chills, fever (100.9), chest pain, dizziness, muscle aches, diarrhea [3 days] | NA |
| G | 46-50 | Female | Asthma<br>Hypertension<br>BMI>30 | No | Pfizer x 2<br>Moderna x 1 | 196 | Ear pain, runny nose, sneezing, no appetite, abdominal pain, fatigue, cough, shortness of breath, sore throat, chills, fever (not measured), chest pain, headache, muscle aches, diarrhea [5 days] | NA |
| H | 36-40 | Female | BMI>25 | No | J&J x 1<br>Moderna x 1 | 184 | Runny nose, sneezing, fatigue, cough, sore throat, chills, fever (101.7), headache, muscle aches [4 days] | NA |
|  |  |  |  |  |  |  | **Median 4 days symptoms (Range 3-8 days)** |  |

Supplemental Table 2. Continued.
| **Rebound (N=3)** |  |  |  |  |  |  |  |  |
| --- | --- | --- | --- | --- | --- | --- | --- | --- |
|  | **Age Range** | **Sex at Birth** | **Comorbidities** | **Prior Infection** | **COVID-19 Vaccines** | **Days from Vaccination** | **Acute Symptoms [Max Duration]** | **Rebound Symptoms** |
| I | 41-45 | Female | Chronic skin condition**<br>BMI>25 | No | Moderna x 2<br>Pfizer x 1 | 189 | Runny nose, fatigue, sore throat, fever (100.2), headache<br>[3 days] | Asymptomatic |
| J | 41-45 | Female | Autoimmune condition on natalizumab**<br>BMI>25 | No | Pfizer x 4 | 68 | Runny nose, sneezing, fatigue, sore throat, dizziness<br>[7 days] | Runny nose, sneezing, fatigue, sore throat, muscle aches<br>[3 days] |
| K | 46-50 | Male | Asthma<br>Diverticulosis | No | Pfizer x 3 | 155 | Joint pain, fatigue, cough, sore throat, dizziness, muscle aches, diarrhea<br>[5 days] | Fatigue, myalgia, runny nose<br>[3-5 days] |
|  |  |  |  |  |  |  | **Median 5 days symptoms (Range 3-7 days)** |  |

\*Note: identifiers A-K are changed from participant study ID and are used for publication only. They are arbitrary and do not correspond to the numbering system used for sequence data presented in Figure 1 and Supplemental Figure 1; this numbering system is also arbitrary.
\*\*Specific condition not described to avoid identification.

**Supplemental Table 3.**
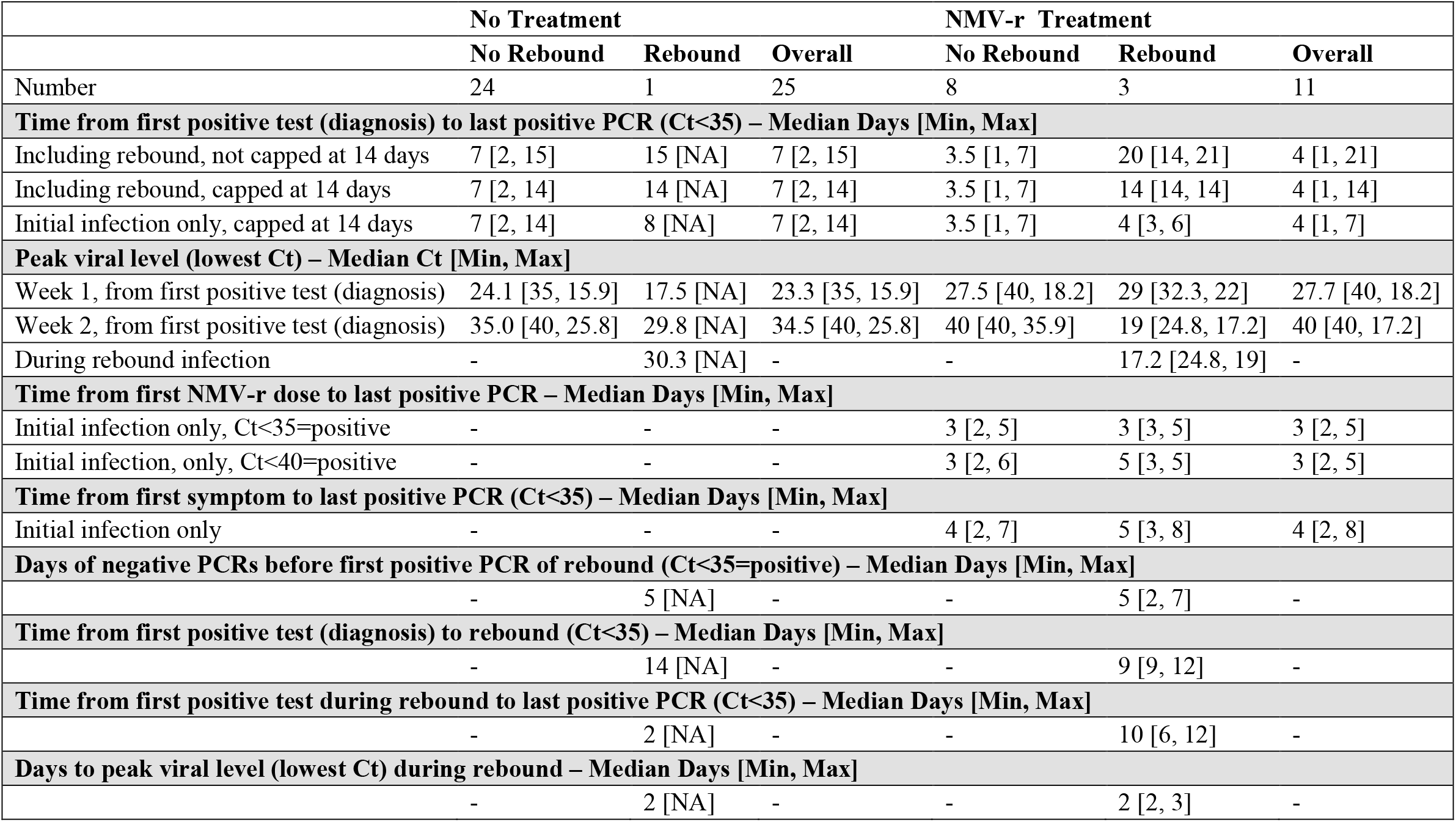
Virologic characteristics of SARS-CoV-2 infection by group

**Supplemental Table 4.**
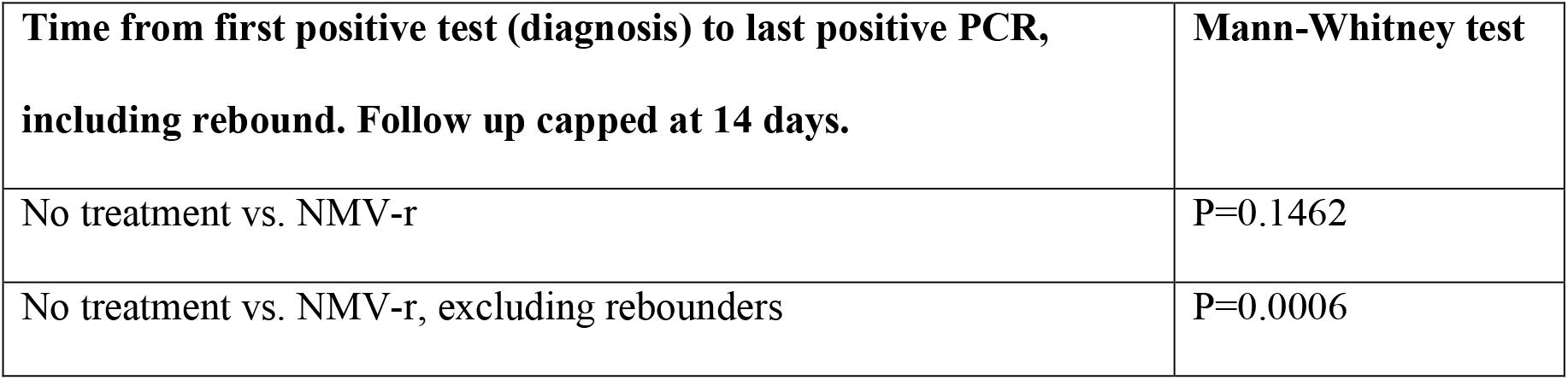
Duration of PCR positivity in no treatment group vs. NMV-r treatment group

## Supplemental Figures

**Supplemental Figure 1.**
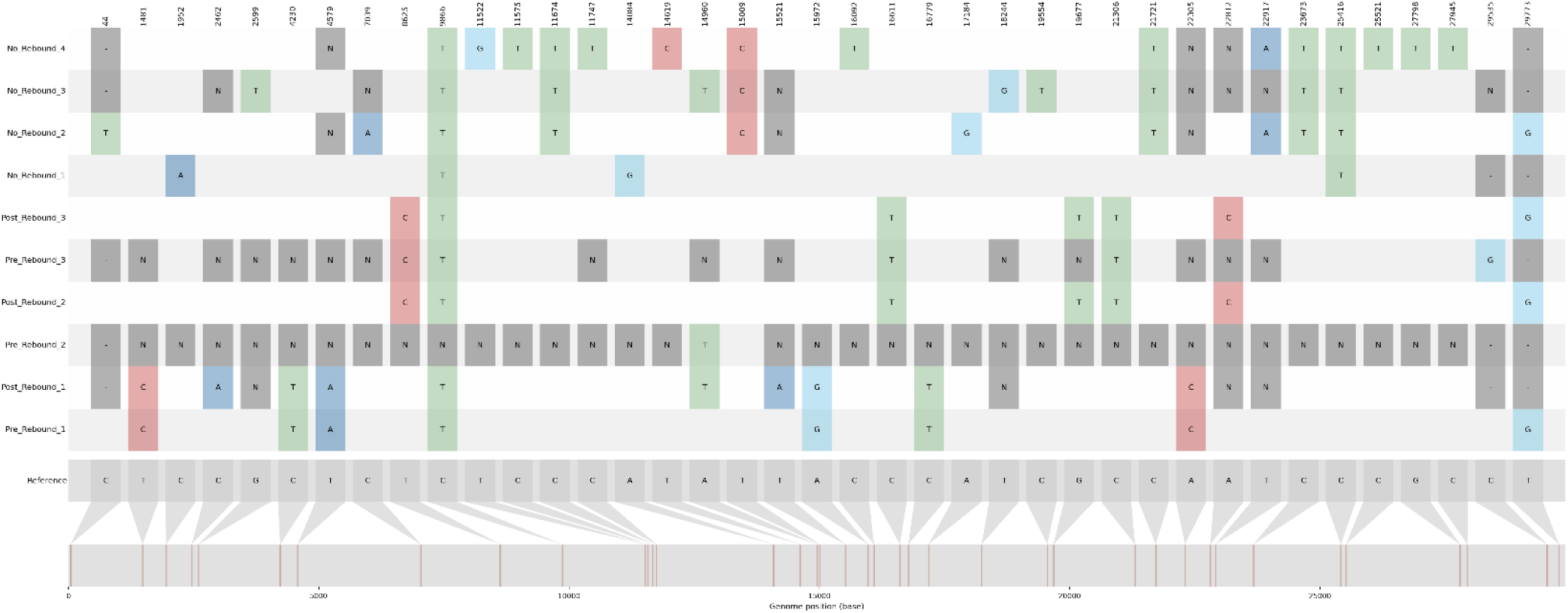
SARS-CoV-2 whole genome sequences, expanded to include all pre-rebound samples. Plot of SARS-CoV-2 whole genome alignment, relative to the BA.2 reference sequence, for viruses in NMV-r treatment group plotted by rebound status (“no rebound”, “pre-rebound”, and “post-rebound”). Single nucleotide variations from reference sequence are noted in color. Pre-rebound samples for Rebound Participants #2 and #3 were of poorer quality (3053 and 20261 unambiguous bases, respectively), but were included to demonstrate the presence of the ancestral allele before, and thus not in response to selective pressures from, drug treatment in all cases where the base was unambiguously resolved. Participant numbering for sequence analysis (e.g. “Rebound Participant #1” etc.) is arbitrary for publication purposes and does not match study identification numbers or the lettering system used in Supplemental Table 2.

